# Body muscle gain and markers of cardiovascular disease susceptibility in young adulthood: prospective cohort study

**DOI:** 10.1101/2020.07.09.20149872

**Authors:** Joshua A. Bell, Kaitlin H. Wade, Linda M. O’Keeffe, David Carslake, Emma E. Vincent, Michael V. Holmes, Nicholas J. Timpson, George Davey Smith

## Abstract

**Background:** The potential benefits of gaining body muscle mass and strength for atherogenic trait levels in young adulthood, and how these compare with the potential harms of gaining body fat, are unknown.

**Methods:** Data were from first-generation offspring of the Avon Longitudinal Study of Parents and Children. Limb lean and total fat mass indices (kg/m^2^) were derived from dual-energy X-ray absorptiometry scans at mean ages 10y, 13y, 18y, and 25y. Maximum handgrip strength was measured using a dynamometer at 12y and 25y, expressed as absolute grip (kg) and relative grip (grip / fat mass index). Linear regression models were used to examine associations of change in standardised measures of these from 10y or 12y to 25y with 228 cardiometabolic traits measured at 25y including metabolomics-derived apolipoprotein-B lipids, glycemic traits, and blood pressure. Changes in lean and fat mass indices across sub-periods of childhood (10y to 13y), adolescence (13y to 18y), and young adulthood (18y to 25y) were also examined with traits at 25y.

**Results:** 3,262 participants (39% male) contributed to analyses. Correlations were positive between changes in lean and fat mass indices, but negative between changes in relative grip and fat mass index. SD-unit gain in limb lean mass index from 10y to 25y was positively associated with atherogenic traits including triglycerides in very-low-density lipoproteins (VLDL). This pattern was limited to lean gain in legs, whereas lean gain in arms was inversely associated with VLDL triglycerides, insulin, glycoprotein acetyls, and others; and was also positively associated with creatinine (a muscle product and positive control). This pattern for arm lean mass index was further specific to gains occurring between 13y and 18y, e.g. −0.13 SD (95% CI = −0.22, −0.04) for VLDL triglycerides. Changes in absolute and relative grip from 12y to 25y were both positively associated with creatinine, but only change in relative grip was also inversely associated with atherogenic traits, e.g. −0.31 SD (95% CI =-0.36, −0.25) for VLDL triglycerides. Change in fat mass index from 10y to 25y was more strongly associated with atherogenic traits including VLDL triglycerides at 0.45 SD (95% CI = 0.39, 0.52); these estimates were directionally consistent across sub-periods with a tendency for larger effect sizes with more recent gains. Associations of lean, grip, and fat indices with traits were more pronounced among males than females.

**Conclusions:** Muscle strengthening is associated with lower atherogenic trait levels in young adulthood, but at a smaller magnitude than unfavourable associations of fat gain. Associations of muscle gain with such traits appear to be smaller and limited to gains occurring in adolescence. These results suggest that body muscle is less robustly associated with markers of cardiovascular disease susceptibility than body fat and may therefore be a lower priority intervention target.

## Background

Cardiovascular diseases remain leading causes of mortality and health service use in Britain (1, 2). Multiple lines of evidence support higher body fat tissue as a strong cause of such diseases, including coronary heart disease (CHD) (3, 4). These harms of body fatness are driven largely by its effects on cardiometabolic intermediates measured in circulation (5) including higher blood pressure, higher apolipoprotein-B-containing lipoproteins, and higher glucose (4, 6-8). Population reductions in body fat remain difficult to achieve, however (9). This reality motivates the direct targeting of intermediate traits and of other metabolically active, and potentially modifiable, body tissues.

Body muscle tissue is metabolically active (10-12) and measurable using dual-energy x-ray absorptiometry (DXA) scans (13) which isolate lean tissue mass from fat and bone. Muscle contraction is expected to be anti-inflammatory and anti-hyperglycaemic (10), but higher total lean mass often shows paradoxically adverse cardiometabolic profiles (14, 15). This may reflect confounding by ectopic fat in abdominal regions (5). Lean mass held specifically within limbs may better isolate skeletal muscle (16) as these compartments correlate most highly with muscle volume measured by magnetic resonance imaging (16, 17). The cardiometabolic benefits of muscle may also be reflected in strength (18) which can be measured indirectly using maximum handgrip tests (19). Prospective observational estimates suggest that higher limb lean mass and stronger grip are both associated with lower CHD risk independent of body mass index (BMI) (19-21), while one Mendelian randomization (MR) study suggested no protection against cardiovascular morbidity or mortality from stronger grip independent of BMI (22). Moreover, although males are known to have higher lean mass and grip strength than females (19, 21, 23), whether these are more favourably associated with cardiometabolic health among males has not been comprehensively examined.

Causality of body muscle for cardiovascular diseases can be interrogated by examining its associations with key intermediates. Higher limb lean mass and stronger grip are associated with lower levels of insulin, glucose, apolipoprotein-B lipids, blood pressure, and inflammatory traits (24-30). These associations for grip strength often appear stronger when expressed as a function of, rather than adjusting for, BMI (25, 28-30). Notably, estimates for limb lean mass and grip strength are also based largely on measures taken on middle-to-older age adults which limits causal inference given the high potential for confounding by ageing-related disease processes, i.e. reverse causation (24, 31, 32). The few existing studies of children or young adults suggest weak associations of stronger grip with total cholesterol, glucose, and blood pressure (33-36); and potentially positive associations of higher limb lean mass with atherogenic lipids, glycemia, and blood pressure (15). The lack of repeated measures of exposure has also limited insight into the potential benefits of muscle gain during periods of growth and development (37, 38). Measuring muscle and strength earlier in life, when subclinical disease is rare, would also naturally reduce reverse causation bias and enable more accurate estimates of their potential benefits for cardiometabolic health.

We aimed to estimate the effects of gaining body muscle mass and strength on markers of cardiovascular disease susceptibility using repeated measures of DXA limb lean mass and grip strength taken across early life in relation to blood pressure and metabolomics-derived lipid, glycemic, and inflammatory traits measured in young adulthood. We examined associations of change in limb lean mass and grip strength from childhood to young adulthood with these traits, and whether these associations differ by stage of body development (childhood, adolescence, young adulthood) and by sex. We also examined associations of change in DXA fat mass index with cardiometabolic traits in the same manner to directly compare the magnitude of potential benefits of gaining body muscle with the potential harms of gaining body fat.

## Methods

### Study population

Data were from offspring participants (Generation (G) 1) of the Avon Longitudinal Study of Parents and Children (ALSPAC), a population-based birth cohort study in which 15,454 pregnant women (G0) with an expected delivery date between 1 April 1991 and 31 December 1992 were recruited from the former Avon county of southwest England (39). Since then, 14,901 G1 offspring alive at one year have been followed repeatedly with questionnaire- and clinic-based assessments (40-42), including an additional 913 G1 children enrolled over the course of the study (43). Written informed consent was provided and ethical approval was obtained from the ALSPAC Law and Ethics Committee and the local research ethics committee. Consent for biological samples has been collected in accordance with the Human Tissue Act (2004). Informed consent for the use of data collected via questionnaires and clinics was obtained from participants following the recommendations of the ALSPAC Ethics and Law Committee at the time. The study website contains details of all available data through a fully searchable data dictionary and variable search tool (http://www.bristol.ac.uk/alspac/researchers/our-data/).

### Assessment of body muscle mass and strength

When aged approximately 10y, 12y, 13y, 18y, and 25y, participants underwent body scanning using a DXA Lunar Prodigy narrow fan beam densitometer, from which lean mass within the trunk, arms, and legs (in kg, excluding fat and bone mass) was estimated. Scans were screened for anomalies, motion, and material artefacts, and were realigned when necessary (44). Limb lean mass was calculated by summing lean mass in arms and legs (left-right body sides combined; trunk mass excluded). On each occasion, height was measured in light clothing without shoes to the nearest 0.1 cm using a Harpenden stadiometer. Limb lean mass index was then calculated using squared-height (kg/m^2^). Separate lean mass indices for arms and legs were calculated for comparison. Fat mass index was also constructed based on total body fat mass from DXA (kg/m^2^).

When aged approximately 12y and 25y, participants underwent handgrip strength testing using a Jamar hand dynamometer. Participants sat in a chair with arms and back supported and were asked to rest their forearms on the arms of the chair with their wrist just over the end of the chair arm (thumb facing upwards with the wrist in a neutral position). Participants were asked to squeeze the device as tightly and for as long as possible, three times in succession using their dominant (writing) hand, to record maximum isometric strength in kg units of force. Maximum grip strength was estimated as the mean of these three measures and is here termed ‘absolute grip’. For comparison, a grip strength value as a function of body fat (termed ‘relative grip’, calculated as grip strength divided by DXA total fat mass index) was also calculated.

### Assessment of cardiometabolic traits

During the 25y clinic, systolic and diastolic blood pressure (SBP and DBP, respectively) were examined twice in succession while seated with the arm supported using an appropriately-sized cuff and a DINAMAP 9301 device. Mean levels of each were used to represent resting SBP and DBP. Fasting blood samples were drawn, from which insulin (mu/l) and C-reactive protein (CRP, mg/l) were quantified using routine clinical chemistry. Proton nuclear magnetic resonance (NMR) spectroscopy as part of a targeted metabolomics platform (45) was also performed using fasting blood samples to quantify 224 traits (145 concentrations mostly in mmol/l, plus 79 ratios) including total lipid, cholesterol, and triglyceride content within various lipoprotein subclasses, apolipoprotein-B concentration, glucose, branched chain and aromatic amino acids, creatinine (a product of skeletal muscle and thus positive control here), inflammatory glycoprotein acetyls, and others.

### Assessment of covariates

The core set of covariates considered as confounders were sex, ethnicity (white vs. non-white), age in months at the time of exposure (lean mass, grip strength, fat mass) assessment, and the highest level of education attained by the participant’s mother as reported shortly after delivery (‘Certificate of Secondary Education (CSE)’; ‘vocational’; ‘O-level’; ‘A-level’; or ‘degree’ using English standards) to indicate socioeconomic position at birth. Smoking at age 25y was recorded via questionnaire and grouped as ‘never smoked an entire cigarette’; ‘smokes less than weekly’; or ‘smokes every week’. Alcohol consumption at 25y was also recorded and grouped as ‘never/monthly/less than monthly’; ‘2 or 4 times per month’; or ‘2 or more times per week’. Puberty timing was estimated through age at peak height velocity based on SuperImposition by Translation And Rotation (SITAR) growth curve modelling of repeated height measures from age 5y to 20y (detailed elsewhere (46)).

### Analytical approach

First, Pearson correlation coefficients were examined between changes in lean and fat mass indices based on ages 10y and 25y; and between changes in grip strength measures, lean mass indices, and fat mass index based on ages 12y and 25y. Correlations were also examined between changes in lean and fat mass indices across their available sub-periods of childhood, adolescence, and young adulthood.

All exposures (lean mass indices, grip strength measures, fat mass index) and outcomes (cardiometabolic traits) were subsequently analysed in standardised units based on z-scores to allow comparability of effect sizes given dissimilar variances between traits and across time points. Because the distributions of exposures differed substantially by sex (**Supplementary Figures 1-5**), as did the distributions of change in exposures based on original units (**Supplementary Figures 6-10**), z-scores for exposures were derived separately within each sex. We then examined prospective associations of change in each lean mass index (total limb, arm, leg) based on a difference score (standardised index at 25y minus standardised index at 10y) with cardiometabolic traits at 25y using linear regression models with robust standard errors. These models adjusted for age, sex, ethnicity, maternal education, the age 10y (baseline) value of the index being assessed, change in the alternative limb compartment, and change in fat mass index. We examined whether association patterns differ by stage of body development by repeating the above analyses of change based on change in lean mass indices from 10y to 13y (childhood), 13y to 18y (adolescence), and 18y to 25y (young adulthood), each in relation to cardiometabolic traits at 25y. Models of change in adolescence were additionally adjusted for age at peak height velocity (puberty timing), while models of change in young adulthood were additionally adjusted for puberty timing plus smoking and alcohol consumption at age 18y. Prospective associations of change in each grip strength measure (absolute and relative) with cardiometabolic traits at 25y were examined using linear regression models with the same adjustment strategy as above (models of change in relative grip were not additionally adjusted for change in fat mass index since this was used to derive relative grip).

For comparison of results for lean mass and grip strength measures, we examined prospective associations of change in fat mass index from 10y to 25y, and over the same sub-periods of 10y to 13y, 13y to 18y, and 18y to 25y, with the same cardiometabolic traits at 25y, using model adjustment strategies described above but with adjustment for limb lean mass index instead of fat mass index. Additionally, we examined whether patterns of association of all exposures with trait outcomes differed substantially by sex by repeating all above analyses among males and females separately.

Analyses were first conducted on unrestricted samples of participants (N varying between traits and across time points) to make full use of measured data. To examine whether results are sensitive to changing sample size, we repeated all models using a complete case sample of 787 participants who had data on every DXA and grip strength measure at every time point, every cardiometabolic trait, and every covariate needed for adjustment.

As recommended (47, 48), we present exact P-values and base our interpretation of results on effect size and precision. Analyses were done using Stata 15.1 (StataCorp, College Station, Texas, USA).

## Results

### Sample characteristics

3,262 participants contributed to analyses; 38.6% were male and 3.9% were of a non-white ethnicity (**Table 1; Figure 1**). Mean (SD) age at peak height velocity was 12.4y (1.2y) overall, ranging from 9.1y to 17.4y. Based on this indicator, 0.4% of participants had entered puberty by the 10y clinic (when lean mass was first assessed), 28.6% had entered puberty by the 12y clinic (when grip strength was first assessed), and all had entered puberty by the 25y clinic. At the time of the 18y clinic, weekly smoking was 15.1%, while alcohol consumption at least bi-weekly was 24.5%. Mean changes in lean mass indices from childhood to young adulthood were generally positive but smaller than positive changes in fat mass index over the same period; all changes involved substantial variability around mean values (**Table 1**). Overall, males experienced more positive change in lean mass indices and grip strength than females, whereas females experienced more positive change in fat mass index than males; these sex differences appeared largest in childhood and adolescence (**Supplementary Figures 6-10**).

**Table 1.** Characteristics of 3,262 ALSPAC G1 offspring eligible for analyses

| **Characteristics** | **N** |  |
| --- | --- | --- |
| Male – % (N) | 3262 | 38.6 (1260) |
| Non-white ethnicity – % (N) | 3262 | 3.9 (126) |
| Highest maternal education is degree – % (N) | 3262 | 20.8 (679) |
| Age (y) at peak height velocity – mean (SD) | 2896 | 12.4 (1.2) |
| Smoking at 18y – % (N) | 2405 |  |
| Never |  | 51.4 (1237) |
| Less than weekly |  | 33.4 (804) |
| Every week |  | 15.1 (364) |
| Alcohol consumption at 18y – % (N) | 2263 |  |
| Never/monthly/less than monthly |  | 27.9 (631) |
| 2 to 4 times per month |  | 47.6 (1078) |
| 2 or more times per week |  | 24.5 (554) |
| ***Changes in muscle and fat indices from childhood to young adulthood – mean (SD)*** |  |  |
| Total limb lean mass index (kg/m<sup>2</sup>), 10y to 25y | 2808 | 1.8 (1.0) |
| Arm lean mass index (kg/m<sup>2</sup>), 10y to 25y | 2808 | 0.6 (0.4) |
| Leg lean mass index (kg/m<sup>2</sup>), 10y to 25y | 2808 | 1.2 (0.7) |
| Absolute grip strength (kg), 12y to 25y | 1739 | -4.2 (4.5) |
| Relative grip strength (kg/kg/m<sup>2</sup>), 12y to 25y | 1677 | -2.5 (2.3) |
| Total fat mass index (kg/m<sup>2</sup>), 10y to 25y | 2808 | 3.6 (3.0) |
| ***Above changes in childhood – mean (SD)*** |  |  |
| Total limb lean mass index (kg/m<sup>2</sup>), 10y to 13y | 2565 | 1.0 (0.6) |
| Arm lean mass index (kg/m<sup>2</sup>), 10y to 13y | 2565 | 0.3 (0.2) |
| Leg lean mass index (kg/m<sup>2</sup>), 10y to 13y | 2565 | 0.7 (0.4) |
| Total fat mass index (kg/m<sup>2</sup>), 10y to 13y | 2565 | 0.9 (1.7) |
| ***Above changes in adolescence – mean (SD)*** |  |  |
| Total limb lean mass index (kg/m<sup>2</sup>), 13y to 18y | 2404 | 0.3 (0.6) |
| Arm lean mass index (kg/m<sup>2</sup>), 13y to 18y | 2404 | 0.2 (0.2) |
| Leg lean mass index (kg/m<sup>2</sup>), 13y to 18y | 2404 | 0.1 (0.5) |
| Total fat mass index (kg/m<sup>2</sup>), 13y to 18y | 2404 | 1.1 (2.0) |
| ***Above changes in young adulthood – mean (SD)*** |  |  |
| Total limb lean mass index (kg/m<sup>2</sup>), 18y to 25y | 2606 | 0.5 (0.7) |
| Arm lean mass index (kg/m<sup>2</sup>), 18y to 25y | 2606 | 0.1 (0.2) |
| Leg lean mass index (kg/m<sup>2</sup>), 18y to 25y | 2606 | 0.5 (0.6) |
| Total fat mass index (kg/m<sup>2</sup>), 18y to 25y | 2606 | 1.6 (2.4) |

Described are those with data on change in at least 1 lean, grip, and fat measure across any occasion, at least 1 cardiometabolic trait at 25y, and basic covariates used for models. Absolute grip strength defined as maximum grip strength (kg). Relative grip strength defined as maximum grip strength (kg) divided by total fat mass index (kg/m<sup>2</sup>).

**Figure 1.**
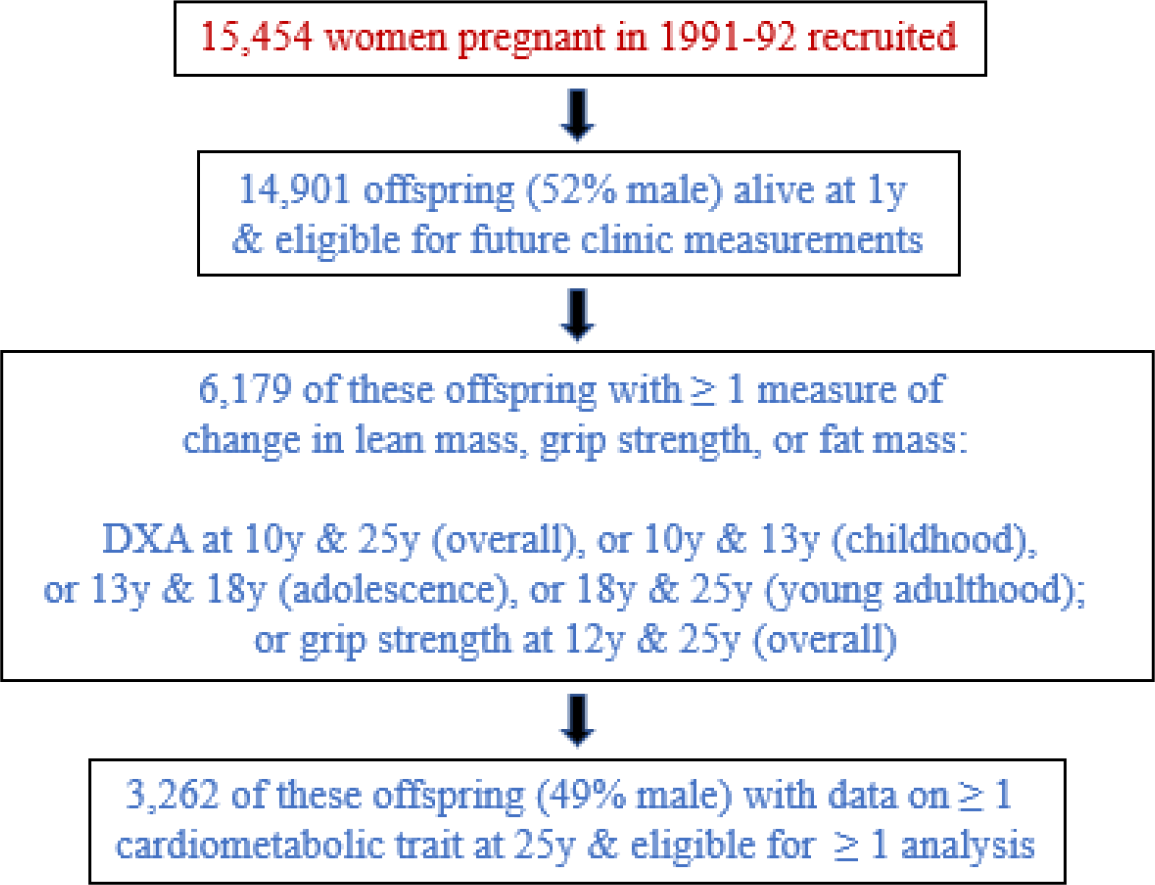
Selection of ALSPAC G1 offspring participants eligible for inclusion in at least one analysis.

Participants who were ineligible for analyses were more likely than eligible participants to be male (54.6%) and to have lower maternal education levels; they also showed a higher prevalence of current smoking and of weekly drinking (**Supplementary Table 1**). Change values of lean and fat mass indices were similar among ineligible vs eligible participants.

### Correlations amongst changes in lean mass indices, grip strength, and fat mass index

Change in limb lean mass index from childhood to young adulthood was positively correlated with change in each limb compartment and absolute grip but was uncorrelated with relative grip. Based on 10y to 25y differences, change in fat mass index was positively correlated with change in limb lean mass index (r = 0.47) and was more positively correlated with change in leg than arm lean mass index (r = 0.45 vs 0.39, respectively; (**Supplementary Tables 2-3**). Based on 12y to 25y differences, change in fat was uncorrelated with change in absolute grip but negatively correlated with change in relative grip. When examining changes across sub-periods, fat mass index was more positively correlated with arm than leg lean mass index in childhood and adolescence (**Supplementary Tables 4-5**), whereas fat change was more positively correlated with change in leg than arm lean mass index in young adulthood (**Supplementary Table 6**).

### Associations of changes in limb lean mass indices with cardiometabolic traits

Evidence was strong for associations of change in limb lean mass index from 10y to 25y (per SD-unit gain, based on standardised index at 25y minus standardised index at 10y, and 10y index as a covariate) with higher creatinine and most atherogenic traits; these were of modest magnitude and mostly in directions assumed to reflect poorer health, e.g. 0.17 SD (95% CI = 0.10, 0.24) higher triglycerides in very-low-density lipoprotein (VLDL) and higher glycoprotein acetyls (**Figure 2; Supplementary Table 7**). When examining gains in arm and leg lean indices separately, only gain in arms was positively associated with creatinine (a known muscle product). The adverse pattern of associations seen across traits with limb lean mass index gain appeared limited to gain in leg lean mass index, whereas gain in arm lean mass index was associated with lower VLDL triglycerides (−0.09 SD, 95% CI = −0.15, − 0.02), and others including lower insulin, glycoprotein acetyls, and DBP, but higher SBP. This pattern for creatinine and atherogenic traits was more pronounced among males than females, i.e. effect sizes were larger (**Supplementary Tables 8-9**).

**Figure 2.**
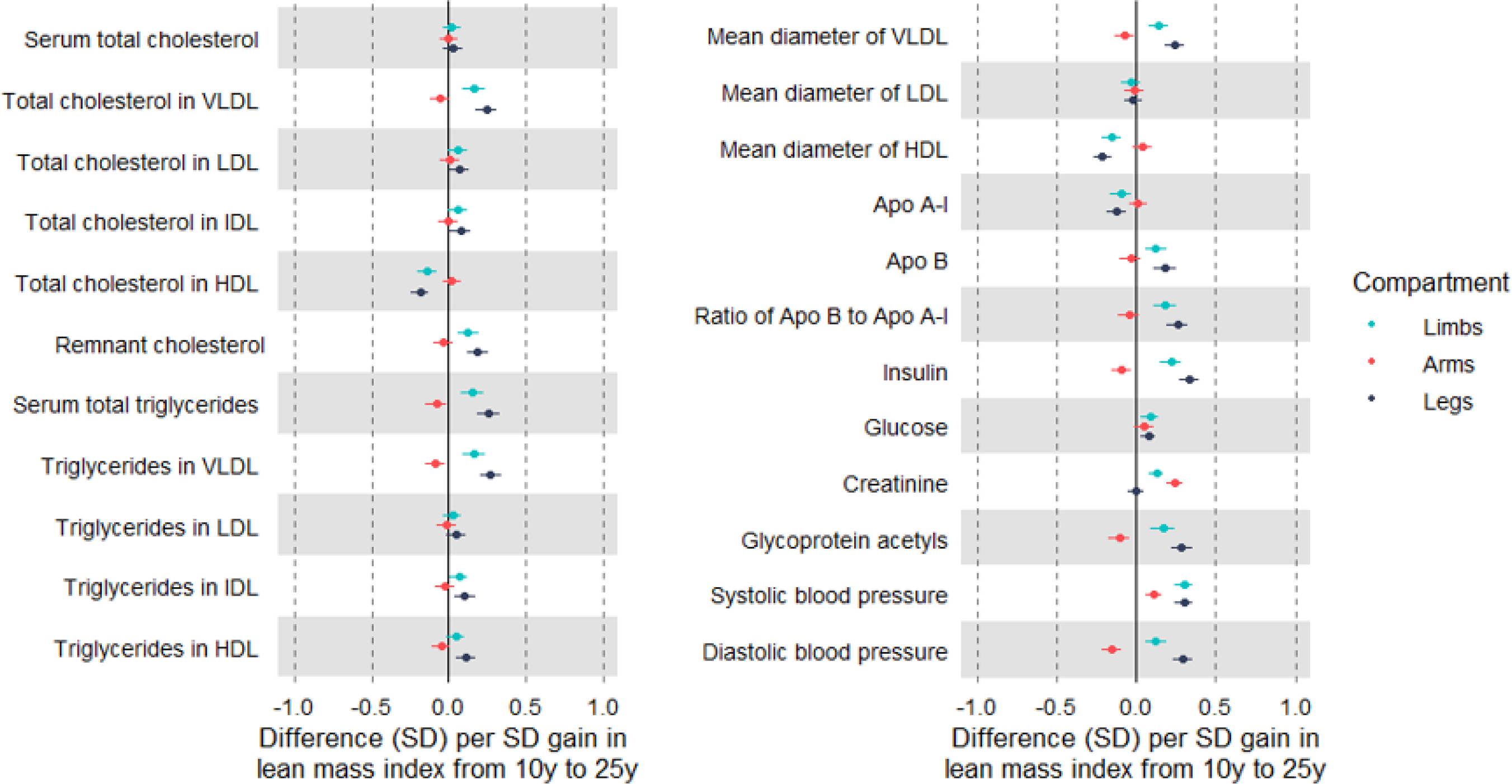
Associations of change in limb lean mass indices from age 10y to 25y with cardiometabolic traits at 25y among ALSPAC G1 offspring. Estimates are beta coefficients and 95% CIs representing SD-unit differences in cardiometabolic traits at 25y per SD-unit gain in lean mass index from 10y to 25y (based on standardised index at 25y minus standardised index at 10y). Models adjusted for age, sex, ethnicity, maternal education, lean mass index at 10y, change in other lean mass index, and change in total fat mass index (N range: 2121 to 2804). Limb lean mass index defined as sum of lean mass in arms and legs (kg) divided by squared-height (m^2^).

Change in limb lean mass index from 10y to 13y was generally associated with higher atherogenic lipids at 25y including total cholesterol in LDL and VLDL triglycerides at 0.19 SD, 95% CI = 0.13, 0.26; positive associations were also apparent with apolipoprotein-B, insulin, glycoprotein acetyls, SBP, and DBP (**Figures 3-4; Supplementary Table 10**). Gain in both limb compartments was positively associated with creatinine, and adverse trait profiles seen with childhood gain in limb lean mass index were reflected in arms as well as legs. Effect sizes tended to be larger among males (**Supplementary Tables 11-12**).

**Figure 3.**
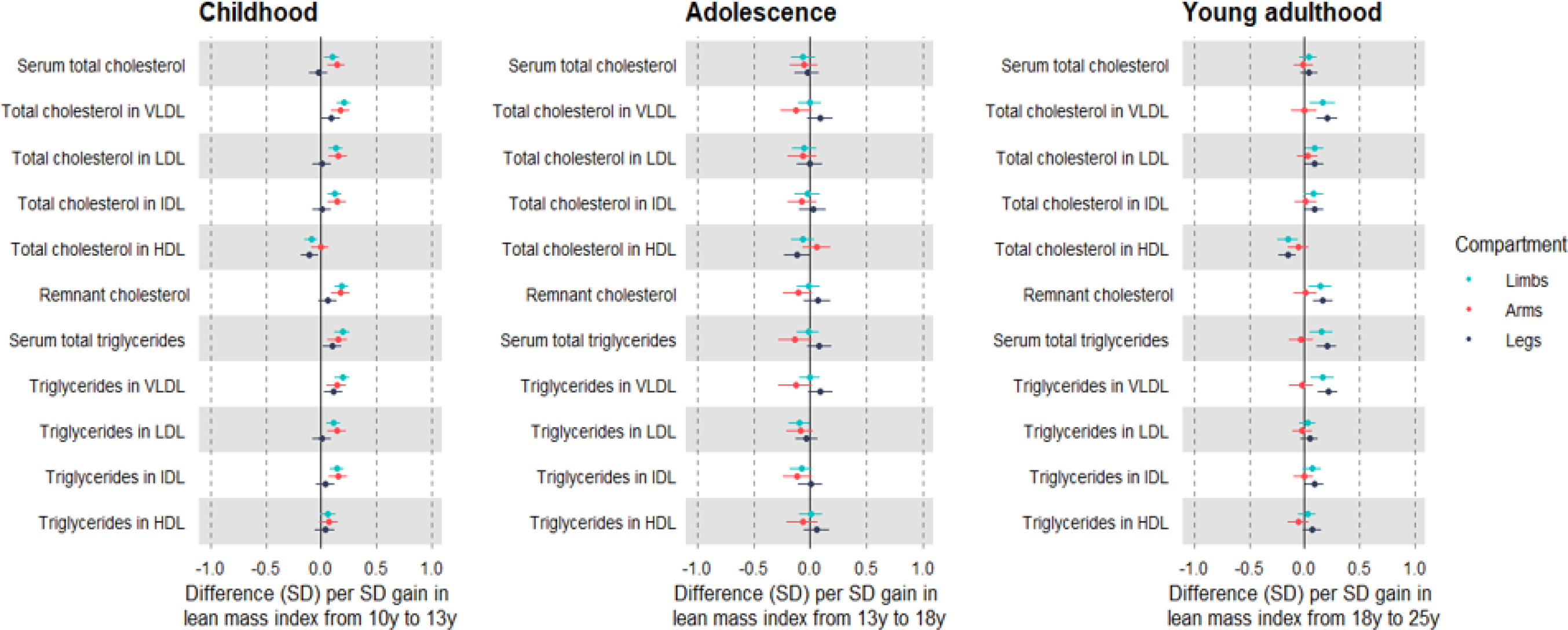
Associations of change in limb lean mass indices across different life stages with lipid traits at 25y among ALSPAC G1 offspring. Estimates are beta coefficients and 95% CIs representing SD-unit differences in cardiometabolic traits at 25y per SD-unit change in lean mass index (based on standardised index at time 2 minus standardised index at time 1). Childhood models are based on change in lean mass index from 10y to 13y and are adjusted for age, sex, ethnicity, maternal education, lean mass index at 10y, change in other lean mass index, and change in total fat mass index (N range: 1926 to 2557). Adolescence models are based on change in lean mass index from 13y to 18y and are adjusted for age, sex, ethnicity, maternal education, puberty timing, change in other lean mass index, and change in total fat mass index (N range: 1747 to 2344). Young adulthood models are based on change in lean mass index from 18y to 25y and are adjusted for age, sex, ethnicity, maternal education, puberty timing, smoking, alcohol, change in other lean mass index, and change in total fat mass index (N range: 1532 to 2036). Limb lean mass index defined as sum of lean mass in arms and legs (kg) divided by squared-height (m^2^).

**Figure 4.**
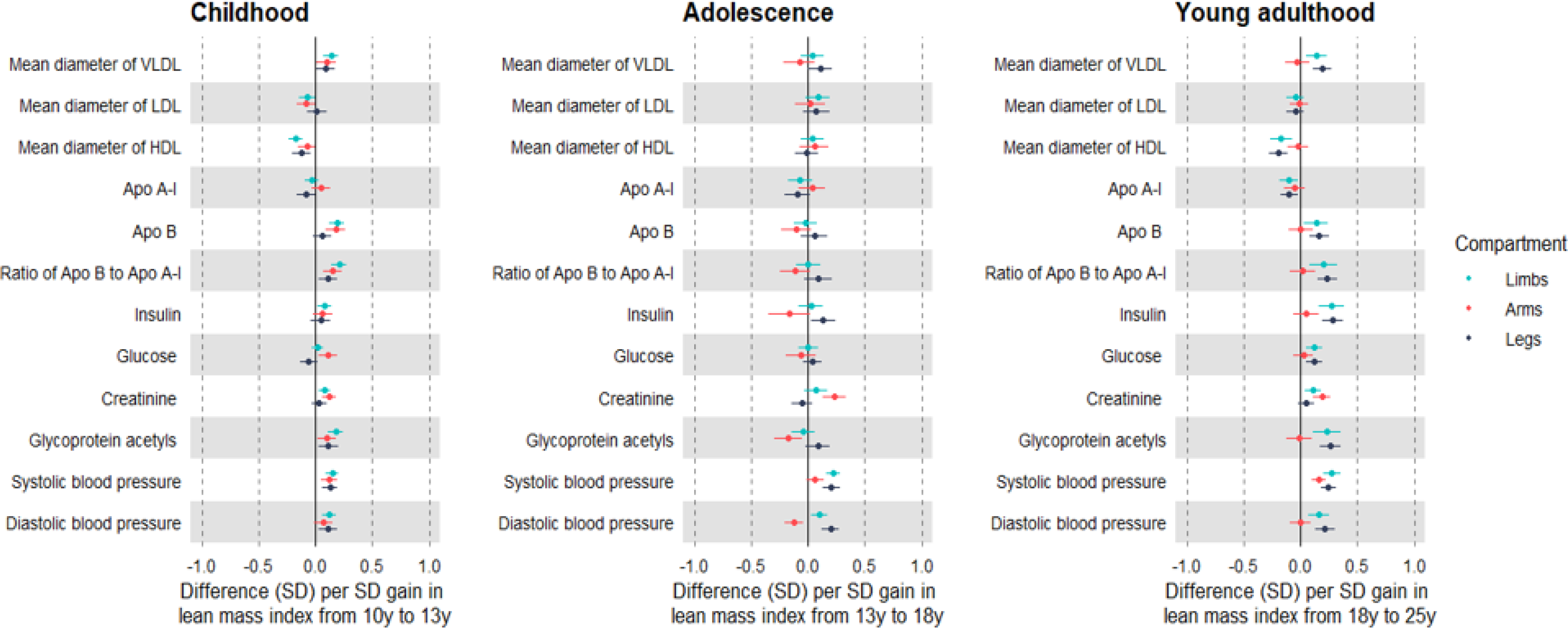
Associations of change in limb lean mass indices across different life stages with lipid, pre-glycemic, inflammatory, and blood pressure traits at 25y among ALSPAC G1 offspring. Estimates are beta coefficients and 95% CIs representing SD-unit differences in cardiometabolic traits at 25y per SD-unit change in lean mass index (based on standardised index at time 2 minus standardised index at time 1). Childhood models are based on change in lean mass index from 10y to 13y and are adjusted for age, sex, ethnicity, maternal education, lean mass index at 10y, change in other lean mass index, and change in total fat mass index (N range: 1926 to 2557). Adolescence models are based on change in lean mass index from 13y to 18y and are adjusted for age, sex, ethnicity, maternal education, puberty timing, change in other lean mass index, and change in total fat mass index (N range: 1747 to 2344). Young adulthood models are based on change in lean mass index from 18y to 25y and are adjusted for age, sex, ethnicity, maternal education, puberty timing, smoking, alcohol, change in other lean mass index, and change in total fat mass index (N range: 1532 to 2036). Limb lean mass index defined as sum of lean mass in arms and legs (kg) divided by squared-height (m^2^).

Change in limb lean mass index from 13y to 18y was generally unassociated with creatinine and atherogenic traits at 25y, apart from weak associations with higher SBP and DBP (**Figures 3-4; Supplementary Table 13**). In contrast, gain in arm lean mass index was positively associated with creatinine and showed inverse associations with atherogenic traits including lower VLDL triglycerides (−0.13 SD, 95% CI = −0.22, −0.04), and lower levels of apolipoprotein-B, insulin, glycoprotein acetyls, and DBP; but higher SBP. These associations were again stronger among males (**Supplementary Tables 14-15**).

Change in limb lean mass index from 18y to 25y was associated with higher creatinine and higher atherogenic lipids at 25y including higher cholesterol and triglycerides in VLDL and LDL; gain was also associated with higher insulin, glycoprotein acetyls, SBP and DBP (**Figures 3-4; Supplementary Table 16**). The positive association with creatinine was again exclusive to arm lean mass index, and gain in arm lean mass index was generally unassociated with atherogenic traits, except for a positive association with DBP. Such null associations were seen both among males and females, despite a strong positive association of gain in arm lean mass index with creatinine among males (**Supplementary Tables 17-18**).

### Associations of change in grip strength with cardiometabolic traits

Change in absolute grip from 12y to 25y (per SD-unit gain) was positively associated with creatinine, but unassociated with atherogenic traits including VLDL triglycerides (**Figure 5; Supplementary Table 19**); this was apparent for males and females (**Supplementary Tables 20-21)**. In contrast, change in relative grip over the same period was positively associated with creatinine and also strongly inversely associated with atherogenic lipid levels, e.g. lower VLDL triglycerides (−0.31 SD, 95% CI = −0.36, −0.25) and lower apolipoprotein-B. Inverse associations were strong between gain in relative grip strength and insulin, glycoprotein acetyls, and blood pressure. These associations of gain in relative grip were strongest among males.

**Figure 5.**
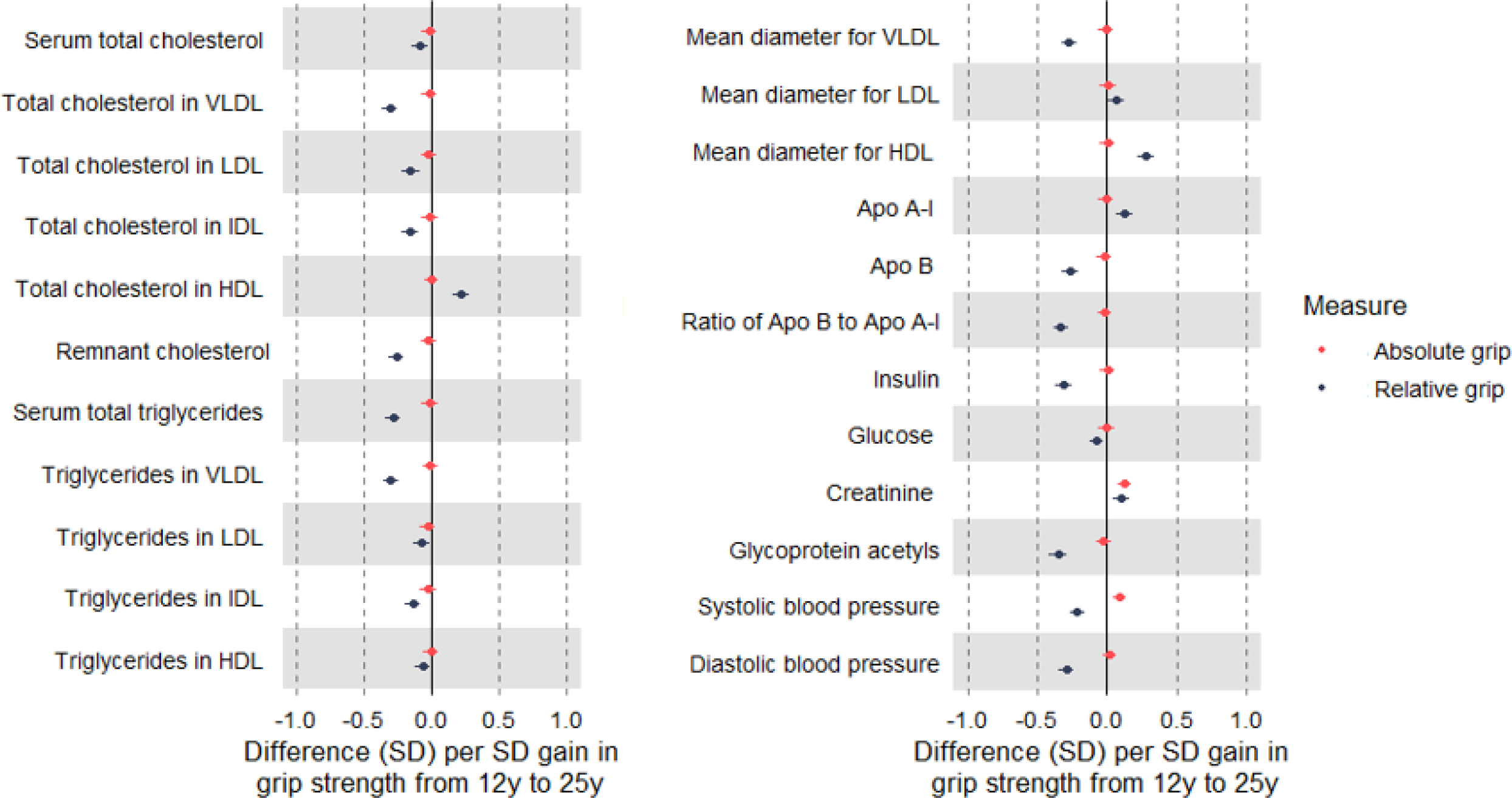
Associations of change in grip strength from age 12y to 25y with cardiometabolic traits at 25y among ALSPAC G1 offspring. Estimates are beta coefficients and 95% CIs representing SD-unit differences in cardiometabolic traits at 25y per SD-unit change in grip strength (based on standardised grip at 25y minus standardised grip at 12y). Models are adjusted for age, sex, ethnicity, maternal education, grip strength at 12y, and change in total fat mass index (except for relative grip models) (N range: 1,246 to 1,678). Absolute grip is based on maximum recorded grip strength of dominant hand (mean of 3 measures, in kg). Relative grip is based on maximum grip strength divided by total fat mass index.

### Associations of change in fat mass index with cardiometabolic traits

Change in total fat mass index (per SD-unit gain) was inversely associated with creatinine and strongly associated with higher atherogenic lipids including VLDL triglycerides (0.45 SD, 95% CI = 0.39, 0.52), LDL cholesterol, and apolipoprotein-B; as well as comparably higher levels of insulin, glycoprotein acetyls, SBP, and DBP (**Figure 6; Supplementary Table 22**). When examining gains over sub-periods in relation to traits at 25y, estimates were found to be directionally consistent across time periods with a tendency for larger effect sizes with more recent gains (i.e. closer to the time of outcome measurement). For example, point estimates for associations of fat gain in childhood, adolescence, and young adulthood with VLDL triglycerides at 25y were 0.10 SD, 0.34 SD, and 0.48 SD, respectively. Associations were consistently more pronounced among males, at about double the magnitude of effect size compared with females (**Supplementary Tables 23-24)**.

**Figure 6.**
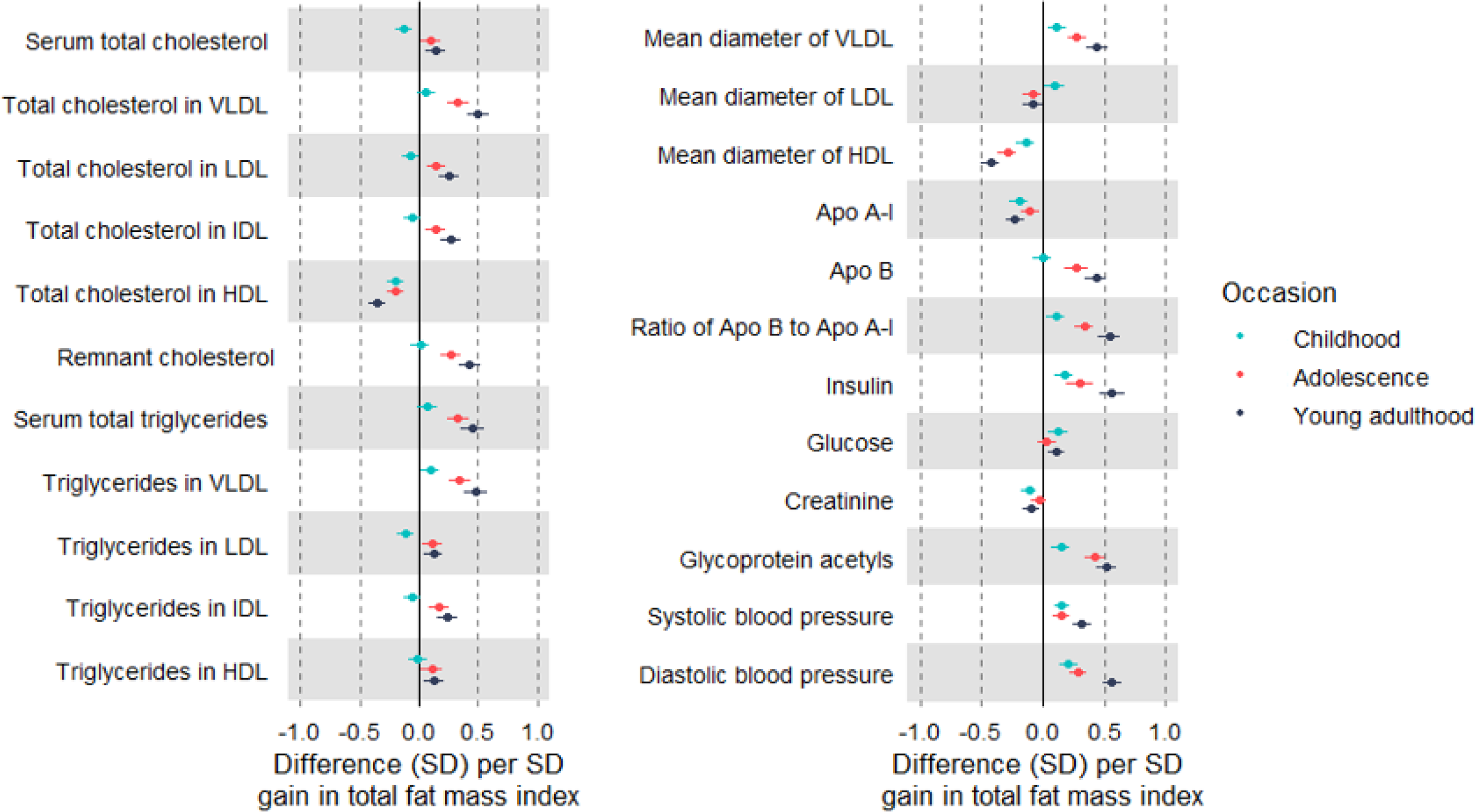
Associations of change in total fat mass index at different life stages with cardiometabolic traits at 25y among ALSPAC G1 offspring. Estimates are beta coefficients and 95% CIs representing SD-unit differences in cardiometabolic traits at 25y per SD-unit gain in total fat mass index in childhood (based on standardised index at 13y minus standardised index at 10y), in adolescence (based on standardised index at 18y minus standardised index at 13y), and young adulthood (based on standardised index at 25y minus standardised index at 18y). Models adjusted for age, sex, ethnicity, maternal education, initial fat mass index, and change in limb lean mass index; models of adolescence additionally adjusted for puberty timing, and models for young adulthood additionally adjusted for puberty timing, smoking, and alcohol at 18y (N range: 2804 to 1532).

Above estimates based on complete-case samples were comparable to those of unrestricted samples (Ns varying) in terms of direction and magnitude of effect size, with expectedly lower precision given smaller sample sizes (**Supplementary Tables 7-24**).

## Discussion

This study aimed to estimate the potential benefits of gaining body muscle mass and strength for markers of cardiovascular disease susceptibility in young adulthood, and how these compare with the potential harms of gaining body fat. We integrated repeated measures of DXA limb lean mass and grip strength starting in childhood with measures of atherogenic traits from targeted metabolomics in young adulthood and examined how the association profiles of change in muscle compare with those of fat. Our results suggest that muscle strengthening is associated with lower atherogenic trait levels in young adulthood, particularly among males. Such associations of gain in muscle mass with traits were smaller in magnitude and limited to gains occurring between ages 13y and 18y. Gaining body fat was more consistently associated with the same atherogenic traits, in unfavourable directions and at larger magnitudes than seen for muscle mass or strength, again particularly among males. Altogether, results suggest that body muscle is less robustly associated with markers of cardiovascular disease susceptibility than body fat and may therefore be a lower priority target for intervention.

The set of cardiometabolic traits considered here includes lipoproteins which are supported through previous ALSPAC analyses as being phenotypic features (i.e. consequences) of greater genetically liability to CHD in mid-adulthood (49). These features of CHD liability, already apparent in childhood (49), most notably include cholesterol and triglycerides within VLDL, LDL, and intermediate-density lipoprotein (IDL) particles, i.e. those containing an apolipoprotein-B molecule which enables lipid-mediated atherosclerosis (50-52). Presently, evidence was very weak for associations of change in limb lean mass index with these lipid types based on prospective assessments. Gain in limb lean mass occurring in adolescence (13y to 18y) was more strongly associated with lower apolipoprotein-B lipids; this appeared to be further limited to adolescent gains within arms, indicating that favourable associations of muscle gain may be sensitive to stage of development and body compartment. These associations appeared to be more pronounced among adolescent males than females. However, the apparent specificity of associations with arms most likely reflects residual confounding of leg lean mass by traces of ectopic fat, rather than genuinely favourable properties of arm versus leg muscle. This is supported by stronger associations of arm lean mass change (versus leg) with higher creatinine (a known product of skeletal muscle), and by weaker correlations of arm lean mass change (versus leg) with total fat mass change. Change in leg lean mass may also better reflect body shape which in turn reflects abdominal fat storage, the compartment that drives effects of total fat gain (14).

Evidence for association of muscle strengthening with atherogenic traits was highly dependent on the measure of grip strength used. Change in grip strength measured in absolute units (not as a proportion of fat mass) was positively associated with creatinine but was generally not associated with any atherogenic trait. In contrast, change in grip strength measured in relative units (as a proportion of fat mass) was also positively associated with creatinine, and was strongly associated with atherogenic traits in directions assumed to indicate better health, e.g. lower apolipoprotein-B lipids, lower fasting insulin, and lower inflammatory levels. The magnitude of these associations of change in relative grip strength was higher than seen for change in arm lean mass index but was generally lower than magnitudes seen for change in fat mass index, particularly among males where evidence tended to be strongest.

The results of other recent ALSPAC analyses of the effects of genetic liability to adult type 2 diabetes on these same cardiometabolic traits measured across early life (53) suggested that phenotypic features of genetic liability, apparent already in childhood, include lower lipids specifically within large and very-large HDL subtypes, higher branched chain amino acid levels, higher glycoprotein acetyls, and (by definition) higher glucose. Although adolescent gain in limb lean mass was favourably associated with glycoprotein acetyls, associations were weak with HDL subtypes, branched chain amino acids, and glucose. Associations were, however, much stronger for higher grip strength as a proportion of fat mass in relation to each of these diabetes-related traits, suggesting that the strength of muscle is more relevant than the absolute mass of muscle for reducing susceptibility to type 2 diabetes in later life. Type 2 diabetes, as clinically defined by high blood glucose, is itself a major risk factor for CHD (54-56). Importantly, the risk of CHD and of CHD mortality conferred by blood glucose is not exclusive to values within the clinically high range; elevated risk is apparent with moderately higher glucose within the clinically normal range (57, 58). This non-clinical risk also accounts for the highest proportion of CHD cases in populations owing to its high prevalence (57, 59), underscoring the need to attend to type 2 diabetes liability in its early subclinical stages.

Resistance-based physical activity, which focuses on strengthening muscle tissue, is supported by the results of several randomised controlled trials (RCTs) among adults with or without metabolic dysfunction as having an effect on reducing SBP and DBP, but as having little-to-no effect on reducing blood glucose, LDL cholesterol, or triglycerides (60, 61). Such trials are typically small (< 100 participants) and have short follow-up durations (< 1 year), but beneficial effects on glycemic and lipid parameters are seen in other RCTs of resistance- and aerobic-based activities among adults with type 2 diabetes (62, 63). Any physical activity biomechanically involves the contraction of some muscle tissue, and habitual physical activity may usefully mark the frequency of such contraction. A large body of prospective observational evidence also supports favourable associations of higher habitual physical activity with better cardiometabolic profiles (64-66); these were also apparent among adolescents in ALSPAC (67), but were generally small in magnitude, at approximately half the size presently seen for body fat. Altogether, evidence seems to indicate that the regular use of muscle tissue matters more than the volume or intentional building-up of muscle tissue for reducing susceptibility to cardiovascular disease.

Whether associations of lean mass gain with susceptibility traits are truly sensitive to stage of body development is uncertain. Presently, the profile of associations of change in arm lean mass index (the limb compartment associated with higher creatinine and thus taken to best reflect muscle) with cardiometabolic traits measured in young adulthood were in directions assumed to be unfavourable to health for gains occurring in childhood (including higher levels of apolipoprotein-B lipids, glucose, and glycoprotein acetyls), favourable for gains occurring in adolescence (including lower levels of these same traits), and null for gains occurring in young adulthood. Adolescence is an active period of tissue growth and development following the onset of puberty (38, 68), but how this may confer exclusive benefits of muscle gain is unclear. The results of several RCTs of resistance-based physical activity among children and adolescents generally support beneficial effects on blood pressure, glycemia, and lipid parameters among both age groups (69); however there is some suggestion of larger benefits with later stages of pubertal maturity (70), possibly reflecting greater modifiability of muscle tissue. Presently, measures of grip strength were not available in adolescence for comparison, but the profile of associations seen for lean mass gain was clearly distinct from the profile seen for fat mass gain which involved associations with atherogenic traits that were in consistent (unfavourable) directions across these same sub-periods, with a tendency for larger effect sizes for more recent gains. Replication of associations observed here across early life in different study samples, with approaches that involve different sources of bias (71), is needed to confirm their robustness.

### Study limitations

The limitations of this study include its observational nature which makes estimates of effect prone to biases from confounding factors which are unmeasured, or which are measured but with error. Major sources of confounding of the effects of body muscle and fat on cardiometabolic trait levels are expected to include subclinical disease processes (reverse causation) and health behaviours such as smoking; however the influence of both of these confounding domains is expected to be lower at younger ages given the natural rarity of existing diseases which are most prevalent at older ages, and the rarity or recency of onset of behaviours such as smoking. Muscle mass was estimated using DXA scans which are less granular than MRI and less able to detect and exclude fat that is stored within muscle compartments, i.e. ectopic fat. We examined correlations amongst change in each limb lean mass compartment and total fat mass which gives some indication as to which lean compartments are most susceptible to confounding by residual fat mass. Grip strength was not measured in adolescence which prevented examination of change in grip across sub-periods. The participants analysed here were born in the early 1990s and were relatively lean and predominantly of white-European ancestry; this limits generalisability to other groups but does reduce potential for confounding by existing disease and population structure. The sample sizes available for analyses were modest, particularly for complete-case analyses; this is a common trade-off of detailed phenotyping and one which reduces the precision of estimates. Replication and refinement of estimates in larger samples of participants is needed. We aimed to analyse traits from targeted metabolomics in a holistic manner; the large scale of analyses, particularly given sex differences, prevented examination of the shape of associations. Associations of change in exposures with outcomes are assumed here to be linear, i.e. negative change (loss) is assumed to be the inverse of positive change (gain); this may not always be the case. Examinations of non-linearity are warranted and would be aided by the prioritisation of cardiometabolic traits with the strongest triangulated evidence of causality for clinically relevant outcomes.

### Conclusions

The results of this prospective cohort study suggest that muscle strengthening is associated with lower atherogenic trait levels in young adulthood, particularly among males. Such associations of gain in muscle mass with traits appear to be smaller in magnitude and limited to gains occurring in adolescence. Gaining body fat was more consistently associated with the same atherogenic traits, in unfavourable directions and at larger magnitudes than seen for muscle mass or strength, again particularly among males. Altogether, results suggest that body muscle is less robustly associated with markers of cardiovascular disease susceptibility than body fat and may therefore be a lower priority target for intervention.

## Data Availability

Individual-level ALSPAC data are available following an application. This process of managed access is detailed at www.bristol.ac.uk/alspac/researchers/access. Cohort details and data descriptions for ALSPAC are publicly available at the same web address.

## Acknowledgements

We are extremely grateful to all the families who took part in this study, the midwives for their help in recruiting them, and the whole ALSPAC team, which includes interviewers, computer and laboratory technicians, clerical workers, research scientists, volunteers, managers, receptionists and nurses.

## Data availability

Individual-level ALSPAC data are available following an application. This process of managed access is detailed at www.bristol.ac.uk/alspac/researchers/access.

Cohort details and data descriptions for ALSPAC are publicly available at the same web address.

## Author contributions

JAB planned the study, conducted analyses, and wrote the first draft. KHW, LMO, DC, EEV, NJT and GDS critically reviewed the intellectual content of manuscript drafts and with JAB approved the final version for submission.

